# Evaluating the Association of Depressive Disorder Symptoms and Moral Injuries in Healthcare Workers during COVID-19 Pandemic

**DOI:** 10.1101/2023.05.20.23290269

**Authors:** Amirhossein Behnampour, Sedigheh Ebrahimi, Amir Bazrafshan, Amirhossein Kamyab, Majid Pakdin, Alireza Ebrahimi

## Abstract

**Background:** Moral injury occurs when negative distressing emotions appear and are suppressed. This could lead to several mental health problems such as depression and post-traumatic stress disorder, and result in long-lasting emotional, behavioral, and social problems. Moral injury, a term more commonly used in war contexts, has come into the spotlight during COVID-19 pandemic. We aimed to evaluate the rate of moral injury and its association with psychological injuries during this healthcare crisis.

**Methods:** We assessed the rates of depression, anxiety, stress, and their association with moral injury among 333 nurses, medical interns, and residents between December 2020 and January 2021. This study was done using validated versions of Depression Anxiety Stress Scales (DASS- and Moral Injury Symptom Scale-Healthcare Professionals (MISS-HP) scores.

**Results:** Totally 333 healthcare professionals participated in this study, mostly aged between 26 to 30 years old. Nearly half of the participants had a clinically significant moral injury. The average scores of anxiety and stress were significantly higher in women. The participants who were single showed higher rates of depression and moral injury than married ones. Moreover, anxiety, stress, depression, and moral injury were higher in nurses than other healthcare professionals. The scarcity of personal protective equipment at the workplace and giving care to patients with end-stage COVID-19 diagnosis were among the factors associated with a higher risk of developing mental health problems.

**Conclusion:** The results of this study showed that anxiety, stress, depression, and moral injury were prevalent among healthcare professionals during COVID-19 pandemic. Also, the rates of anxiety, stress, and depression were associated with moral injuries.

## 1. Background

Moral injury occurs when negative emotions such as anger, guilt, shame, and disgust about the world, self, and others appear and are suppressed [1]. This term originated in the war contexts and is considered to result from activities, or lack of them, which could violate one’s ethical or moral code [2]. Commonly, moral injury is not categorized as a psychological disorder; however, it can lead to depression, post-traumatic stress disorder (PTSD), inability to function well, and even suicidal ideation [3]. The impacts of moral injury on individuals are not always recognizable, but the psychological damages caused by it may result in long-lasting emotional, behavioral, and social problems such as anger, depression, anxiety, burnout, and compassion fatigue [4].

Even after about four years since the first case of severe acute respiratory syndrome coronavirus 2 (SARS-CoV-2) was diagnosed, the disease still leads to morbidity and mortality around the world [5, 6]. The social, political, and economic crises caused by coronavirus disease (COVID-19) could be compared to the 1918 influenza pandemic, as it threatened and impacted millions of lives globally [7, 8]. In addition to the socio-economic impacts, COVID-19 pandemic devastatingly affected the mental health of the general population; humans were shocked by the images that once had seemed like a horror tale, such as mass graves that occasionally happened in some countries [9].

During the pandemic, healthcare professionals (HCPs) were relatively more at risk of developing mental health problems because they had to execute medical interventions in complex circumstances, for example, when the appropriate resources were limited [3, 10]. In some cases, they had to isolate themselves and live far away from their families in order to decrease the risk of spreading the contagion to their relatives. Furthermore, HCPs were required to work long hours under extreme pressure, which, in turn, increased the chance of mental problems and moral injuries [11].

HCPs were at the frontlines, facing various stressors such as giving care to end-stage patients, being forced to wear masks and face shields for long hours, and fearing to spread the virus to their relatives. These stressors might result in psychological injuries, which subsequently could lead to lower quality of care for patients and decrease health status of general population [12]. In this study, we aimed to assess the prevalence of moral injuries and depressive disorders during COVID-19 pandemic. We also aimed to examine the association of depressive disorder symptoms and moral injury during this healthcare crisis.

## 2. Methods

### 2.1. Study Design and Population

This study was a cross-sectional survey-based study conducted on 333 HCPs between December 2020 and January 2021 in educational hospitals affiliated with Shiraz University of Medical Sciences, Shiraz, Iran. The sampling method used was chain-referral sampling, which began from a group of nurses, medical interns, and residents that were invited to fill in the questionnaire. Afterwards, the initial group of participants were asked to distribute the online questionnaire among their colleagues.

### 2.2. Questionnaires

The HCPs were invited to participate in the study via social media platforms, and an online platform was used to gather the surveys. The questionnaire had three main components designed to gather the demographic characteristics, Depression Anxiety Stress Scales (DASS-21) and Moral Injury Symptom Scale-Healthcare Professionals version (MISS-HP) scores (**Suppl. 1-5)**.

### 2.3. DASS-21 Questionnaire

DASS-21 is a self-reported screening instrument used for general population and patients, providing measures of depression, anxiety, and stress, independently [13]. The depression subscale investigates dysphoria, hopelessness, devaluation of life, self-depreciation, lack of interest, anhedonia, and inertia. The anxiety subscale measures autonomic arousal, musculoskeletal effects, situational anxiety, and subjective experience of anxious effect. Stress subscale evaluates difficulty relaxing, nervous arousal, being easily upset or agitated, and irritability. Each subscale has seven items rated on a 4-point Likert scale (0-3), total score averaging from 0 to 63, and a higher score indicates higher depression, anxiety, and stress levels. The severity levels of symptoms could be indicated by the score in each domain [14]. The cross-cultural validity of the DASS-21-Persian as a measure of related symptoms among the general population in Iran has been evaluated in previous studies [15]. DASS-21-Persian internal consistency was calculated using Cronbach’s alpha (α) which was an acceptable rate of 0.94 [16].

### 2.4. MISS-HP Questionnaire

MISS-HP is a 10-item questionnaire that covers various dimensions of moral injury. Each item has response choices ranging on a 10-point Likert scale from 1 (“absolutely disagree”) to 10 (“absolutely agree”). A total score ranges from 10 to 100, and higher scores are an indicator of more significant moral injury [17]. In order to use MISS-HP in our study, it was translated into the Persian version, and the subsequent questionnaire was validated by a pilot study.

Receiver operator curve (ROC) was conducted to identify the proper cutoff on the MISS-HP-Persian, which could be used to diagnose healthcare professionals with symptoms of clinically significant moral injuries (**Suppl. 6)**. The optimum score for a cutoff point on the MISS-HP-Persian to identify healthcare professionals with clinically significant moral injury symptoms was 42 or higher, according to Youden’s index from the ROC. With this cutoff point determination, the sensitivity and specificity for detecting moral injury symptoms accompanied by significant functional impairment were 70% and 63%, respectively (positive predictive value: 53%, negative predictive value: 79%).

Content validity assessment of the MISS-HP-Persian was evaluated qualitatively and quantitatively. To assess the qualitative content validity, the MISS-HP-Persian was given to thirty-five experts who were asked to leave a comment on the scale items regarding item allocation, wording, scaling, and grammar. The MISS-HP-Persian final version was created after analyzing and applying their remarks. For quantitative content validity, we used Content Validity Ratio (CVR) defining the necessity of items and Content Validity Index (CVI) defining the relevance of items (**Suppl. 7**.). The experts were asked to rate the need of the MISS-HP items based on a 3-point Likert-type scale consisting of “necessary”, “useful but not necessary”, and “not necessary”. Additionally, they were asked to rate the relevance of items on a 4-point Likert-type scale. The method of content validation is based on previous publications [18]. The questionnaire’s internal consistency was also calculated using Cronbach’s alpha (α) as an acceptable rate of 0.73 [16].

### 2.5. Statistical analysis

All statistical analysis was performed using SPSS version 24.0. Mean and standard deviation (SD) was used to present continuous variables, while categorical variables were shown as numbers and percentage. Univariate analysis was done using independent *t*-test, and Pearson and Spearman correlation coefficient test. Multivariable analysis was also done after checking variance inflation factor (VIF) and collinearity using One-Way ANOVA (Analysis of Variance). P values < 0.05 was considered as the significance level.

### 2.6. Ethical Considerations

This study protocol was approved by the Medical Ethics Committee of Shiraz University of Medical Sciences (Reg. no. 21417). The participants were asked to fill out an informed consent that was provided at the beginning of the questionnaire. The anonymity and secrecy of the participants were guaranteed.

## 3. Results

A total number of 333 healthcare professionals participated in this study, mostly in the 26 to 30 years age group (n=147, 44.1%). The study population consisted of 107 nurses (32.1%), 125 medical interns (37.5%), and 101 medical residents (30.3%). Among participants, 195 were female (58.6%), and 138 were male (41.1%). More than half of the responders were single (61.6%). **Table 1** represents the occupational and demographic characteristics of the respondents.

**Table 1.** Occupational and demographic characteristics of participants (n=333)

| Variable | N (%) |
| --- | --- |
| **Sex, n (%)** |  |
| Female | 195 (58.6) |
| Male | 138 (41.4) |
| **Age group** |  |
| 21-25 years old | 106 (31.8) |
| 25-30 years old | 147 (44.1) |
| >30 years old | 80 (24.0) |
| **Marital status** |  |
| Single | 205 (61.6) |
| Married | 128 (38.4) |
| **Professional category** |  |
| **Medical Interns** | 125 (37.5) |
| **Medical Residents** | 101 (30.3) |
| **Nurses** | 107 (32.1) |
| **Years in profession** |  |
| 1-3 year | 200 (60.1) |
| 4-7 year | 94 (28.2) |
| >7 year | 39 (11.7) |

**Table 2** demonstrates the results of DASS-21 Persian questionnaire distributed among the participants. Moderate to extremely severe anxiety was detected in 23 (16.7%) of male participants. The rate was 52 (37.7%) and 35 (25.4%) for stress and depression in male participants, respectively. Meanwhile, anxiety, stress, and depression were diagnosed in 45 (23.1%), 78 (40%), and 56 (28.7%) female participants, respectively.

**Table 2.** The severity of depression, anxiety, and stress symptoms in participants

|  |  |  |  |
| --- | --- | --- | --- |
| Severity | Males (N=138) | Females (N=195) | Overall (N=333) |
| **Depression** |  |  |  |
| Normal | 87 (63%) | 117 (60%) | 204 (61.3%) |
| Mild | 16 (11.6%) | 22 (11.3%) | 38 (11.4%) |
| Moderate | 19 (13.6%) | 36 (18.5%) | 55 (16.5%) |
| Severe | 7 (5.1%) | 17 (8.7%) | 24 (7.2%) |
| Extremely Severe | 9 (6.5%) | 3 (1.5%) | 12 (3.6%) |
| **Anxiety** |  |  |  |
| Normal | 92 (66.7%) | 135 (69.2%) | 227 (68.2%) |
| Mild | 23 (16.7%) | 15 (7.7%) | 38 (11.4%) |
| Moderate | 15 (10.9%) | 23 (11.8%) | 38 (11.4%) |
| Severe | 4 (2.9%) | 14 (7.2%) | 18 (5.4%) |
| Extremely Severe | 4 (2.9%) | 8 (4.1%) | 12 (3.6%) |
| **Stress** |  |  |  |
| Normal | 67 (48.6%) | 85 (43.6%) | 152 (45.6%) |
| Mild | 19 (13.8%) | 32 (16.4%) | 51 (15.3%) |
| Moderate | 31 (22.5%) | 39 (20%) | 70 (21.0%) |
| Severe | 13 (9.4%) | 27 (13.8%) | 40 (12.0%) |
| Extremely Severe | 8 (5.8%) | 12 (6.2%) | 20 (6.0%) |

As presented in **Table 3**, the average scores of anxiety and stress were significantly higher in women (p-value<0.05). However, the mean value of depression score in the two gender groups was not significantly different (p-value=0.063). The average score of depression had a significant difference between single and married participants (p-value=0.007); while, anxiety and stress had no significant relationship with marital status (p-value>0.05). The average score of depression, anxiety, and stress among the participants had a significant difference according to the type of profession, and nurses experienced more depression, anxiety, and stress than other professionals (p-value<0.05). Our findings also indicated that the lack of personal protective equipment (PPE) at the workplace increases depression, anxiety, and stress (p-value<0.05). Moreover, the average score of depression and stress has a significant relationship with the work experience with the death of COVID-19 patients (p-value<0.05) (**Table 3**).

**Table 3.**
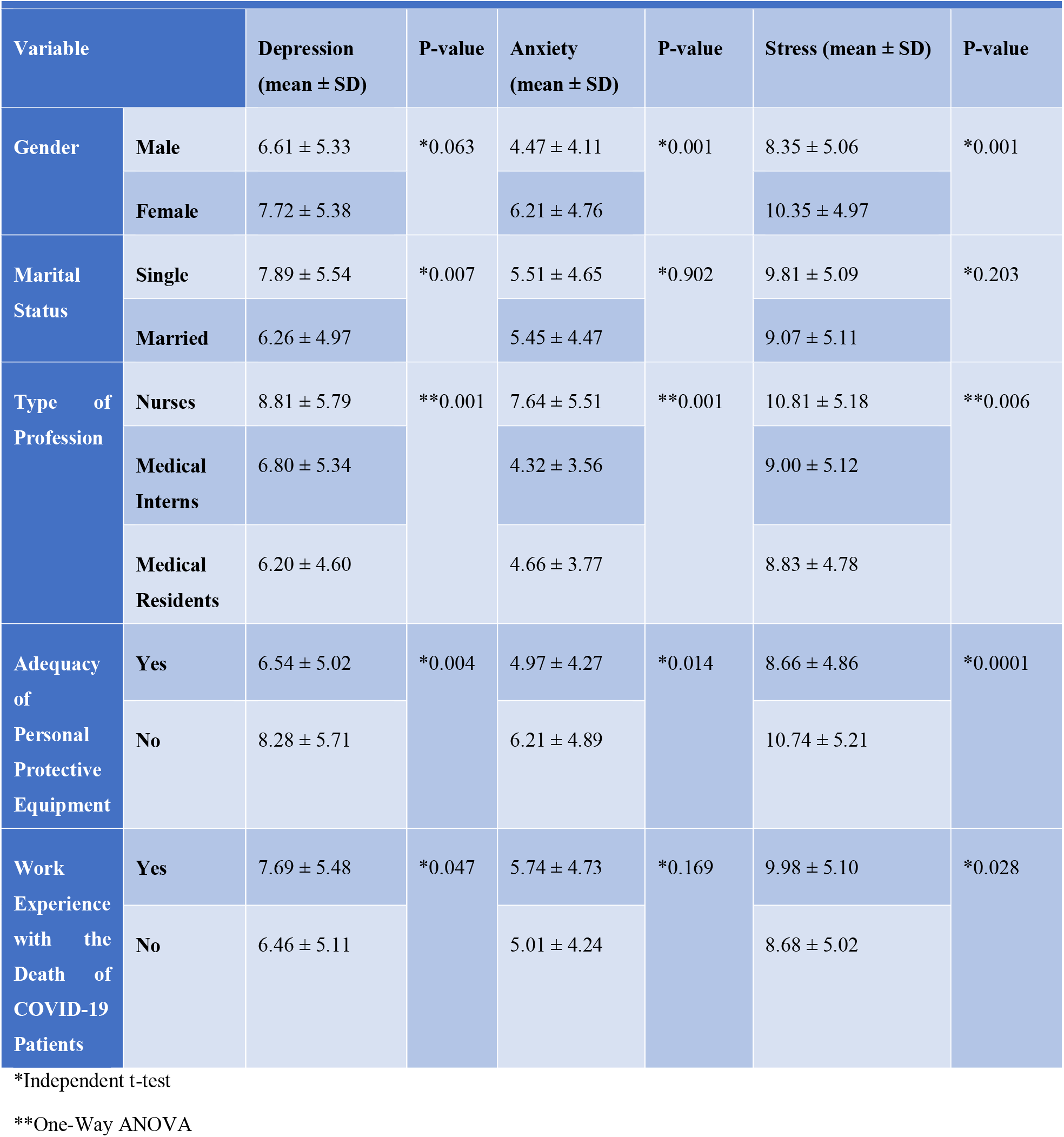
The relationship between demographic variables and depression, anxiety, and stress

According to the results of this study, almost half of the participants had clinically significant moral injury (**Table 4**). Based on our findings, female participants had higher rates of moral injury than their male counterparts (p-value=0.018) (**Table 5**). As shown in **Table 5**, single participants suffered more moral injury than married ones (p-value=0.044). Our results also demonstrated that moral injury was significantly higher in nurses compared to other health care professionals (p-value=0.007). Furthermore, moral injury was significantly correlated with the inadequacy of PPE at the workplace as well (p-value=0.0001) (**Table 5**). We have also observed that moral injury outcomes were positively correlated with depression, anxiety, and stress scores (p-value<0.05) (**Table 6**).

**Table 4.** Moral injury outcome

3 \*Independent t-test 4 \*\*One-Way ANOVA
| Table 4. Moral injury outcome |  |
| --- | --- |
| Moral injury outcomes | n ( %) or Mean (SD) |
| Score obtained in MISS-HP | 41.51 (12.96) |
| Clinically significant moral injury | 165 (49.6) |
| Non-clinically significant moral injury | 168 (50.4) |

**Table 5.** The relationship between demographic variables and moral injury Variable Moral Injury (mean ± SD) P-value

| Table 5. The relationship between demographic variables and moral injury |  |  |  |
| --- | --- | --- | --- |
| Variable | | Moral Injury (mean ± SD) | P-value |
| Gender | Male | 39.51 ± 11.81 | *0.018 |
| | Female | 42.92 ± 13.56 | |
| Marital Status | Single | 42.63 ± 12.86 | *0.044 |
| | Married | 39.70 ± 12.95 | |
| Type of Profession | Nurses | 44.01 ± 13.33 | **0.007 |
| | Medical Interns | 41.88 ± 11.86 | |
| | Medical Residents | 38.39 ± 13.34 | |
| Adequacy of Personal Protective Equipment | Yes | 39.26 ± 12.75 | *0.0001 |
| | No | 44.64 ± 12.63 | |
| Work Experience with the Death of COVID-19 Patients | Yes | 42.37 ± 13.18 | *0.095 |
| | No | 39.87 ± 12.42 | |

\*Independent t-test
\*\*One-Way ANOVA

**Table 6.** Correlation of depression, anxiety symptoms and stress with moral injury

| Disorder | Correlation with moral injury | *P value |
| --- | --- | --- |
| Depression | 0.471 | 0.001 |
| Anxiety | 0.430 | 0.001 |
| Stress | 0.464 | 0.001 |

\* Pearson correlation coefficient test

## 4. Discussion

This study assessed the prevalence of moral injuries, stress, anxiety, and depression among HCPs during the COVID-19 pandemic. The prevalence of anxiety in the study population was 20.4%, consistent with previous studies’ findings in Italy, China, and India [19-21]. In our study, the prevalence of depression was 27.3%, comparable to the findings of Rossi et al. study [19]. Other studies also observed increased level of anxiety and depression among HCPs at the frontlines of outbreaks, for example, among volunteers treating patients with Ebola in West Africa [22]. In our study, 39.1% of participants had moderate to highly severe stress symptoms, which is significantly higher than the corresponding rate found by Lenzo *et al*. using the same assessing tool [23]. These variations could be explained by the phase of the outbreak when studies were conducted. As we conducted our study between the third and fourth peaks of the pandemic in Iran, higher rates might be due to increased burden on healthcare systems during the peak phase of the outbreak. The duration and degree of exposure to COVID-19 patients, type of study, characteristics of the population participating in the study, assessing tool, and sociocultural differences are among other factors which could lead to variation in the findings of different studies.

Our results showed that about one out of two HCPs was suffering from moral injury during COVID-19 pandemic, and also 49.1 percent of all the participants had clinically significant moral injury. These findings are consistent with previous evidence showing a major proportion of HCPs are at risk of moral injury during the pandemic [24].

In this study, although female participants were more at risk of developing anxiety, stress, and moral injury, no significant difference was found in the levels of depression in different genders. Previously, Wang *et al*. reported that female HCPs were more vulnerable to moral damage during COVID-19 pandemic [24]. Additionally, previous investigations showed a higher prevalence of anxiety, stress and depression in female HCPs, during the pandemic [25, 26]. The higher levels of these psychological distresses among women were also seen in general population, and it is assumed that women are more vulnerable to develop anxiety, stress, and depression than men [27-29].

The results of our study showed that the level of depression and moral injury was significantly lower in the married participants. In line with our results, in Wang *et. al* reported lower rates of in married healthcare workers [24]. Similarly, married individuals showed higher levels of resilience against anxiety, stress, and depression during COVID-19 pandemic [25]. Previous studies also considered this factor as a risk for developing depression, caused due to several social and psychological reasons [30-32].

The results obtained from the present study showed that there was a significant relationship between the type of profession and the level of anxiety, stress, depression and moral injury. In consistent with previous published evidence, our study indicated that nurses suffered from anxiety, stress, depression and moral injury more than the medical interns and residents [24]. This difference might be because they spend more hours providing medical services for patients with COVID-19 than medical interns and residents. Xiao *et al*. also observed higher prevalence of anxiety and depression among nurses, however, the difference was not significant between different groups of HCPs [33].

The lack of PPE in the workplace increased the participants’ level of anxiety, stress, depression, and moral damage. This finding is consistent with previous reports that indicated the inadequacy of PPE significantly increased the level of anxiety and depression among HCPs [33]. Adequate provide of PPE against infection was also related to mitigated anxiety and depression in HCPs, in other studies [34]. The fear of contracting the virus and spreading it due to the lack of protective equipment might be the cause of this heightened rates of anxiety and depression.

We also observed that HCPs who experienced working with end-stage patients fighting COVID-19 had higher levels of stress and depression. In this regard, previous studies have found a strong correlation between death anxiety and mental disorders such as depression [35]. Unsurprisingly, the physicians might sympathize with these patients, subsequently leading to feelings of fear, grieve, and depression.

Our findings showed a significant correlation between self-reported depressive symptoms and moral injury in the study population. This finding was consistent with a recent study evaluating an association between psychiatric symptoms and moral injury among HCPs during COVID-19 pandemic [36]. Previous studies also demonstrated an association between psychopathology and moral injury among military veterans [2, 37-41]. Furthermore, trauma-related stress has been highlighted in a recent review evaluating traumatic responses among HCPs during the COVID-19 outbreak [42]. Consistent with the findings of Asalem et al., we found that the effects of COVID-19 pandemic on HCPs simulate the impacts following a traumatic event [36]. This concern was also evaluated by Ahmed *et al*. regarding Ebola and Swine flu, and noticed a rising apprehension towards these outbreaks [43]. Other publications have also highlighted the importance of moral injury among HCPs during the pandemic [44-47].

We found that the prevalence of anxiety, stress, depression, and moral injury was considerably high during COVID-19 pandemic. Also, the rates of anxiety, stress, and depression were associated with moral injury scores. Several factors might be related to this increased rates such as inadequacy of PPE. However, more studies need to be done to find the reasons that might lead to increased risk of developing these responses. Healthcare policymakers and other stakeholders also need to implement strategies to mitigate this concern. This might be achieved by providing adequate sufficient resources (e.g. PPE) and support (e.g. mental health screening) for the healthcare providers during healthcare crises. Providing a supportive environment is of paramount importance [48, 49].

There are several limitations to our study: While the participants were from all section of the hospitals, there is a possibility of selection bias, as our study was a web-based survey. Findings are limited to the population that sample size was derived from, who may not represent the whole HCPs in other regions of the world. Furthermore, our assessment was based on self-report questionnaires which do not meet the accuracy of a formal diagnostic interview.

## 5. Conclusion

The results of this study showed that anxiety, stress, depression and moral injury are prevalent among HCPs during COVID-19 pandemic, and moral injuries associated with the level of anxiety, stress and depression among the participants. Female HCPs showed higher prevalence of anxiety and stress during the pandemic than their male counterparts. Additionally, being married could be a protective factor against depression and moral injury. Different levels of anxiety, stress, depression, and moral injuries were observed in participants with different positions, and nurses were more vulnerable to developing these problems. Having encountered with end-stage patients and inadequacy of resources such as PPE were other factors leading to increased anxiety, stress, depression, and moral injury rates during the crisis.

A growing body of evidence suggests that the prevalence of mental health disorders, including anxiety, depression, and stress has increased among HCPs since COVID-19 eruption. The present study has also highlighted the increased prevalence and significance of moral injuries in this population. These problems could lead to decreased quality of care received by the general public, subsequently resulting in lowered public health [50]. Further research is needed to find proper policies that could be implemented to reduce the consequences of this high amount of moral injuries among HCPs. Researchers and policymakers should take reasonable steps to determine the severity and scope of the problem and try to prevent and resolve this problem in future pandemics, which can be even more widespread and fatal. Regular therapy visits, giving reassurance to HCPs by expert psychologists, and mindfulness training are some examples that have been successful in reducing psychological complications in other pandemics [46, 51, 52].

## Data Availability

All data produced in the present study are available upon reasonable request to the corresponding author.

## 7. Tables

## 8. Supplementary

### 8.1. Suppl 1. Demographic Information Questionnaire

1. Age:
2. Gender:
3. Marital Status:
4. Type of profession: □Nurse □ Medical Intern □ Medical Resident
5. Duration of service as a health care professional:
6. Are you provided with sufficient personal protective equipment at workplace?
7. Have you ever worked in the wards where patients with COVID-19 were hospitalized?
8. How long have you been working in the wards where patients with COVID-19 were hospitalized?
9. Have you or your relatives been infected with COVID-19?
10. If you or your relatives have ever been infected or been hospitalized because of COVID-19?
11. Have you ever had a close work experience with the COVID-19 patients?

### 8.2. Suppl 2. DASS-21

### 8.3. Suppl 3. DASS Severity Ratings

### 8.4. Suppl 4. DASS-21 Domain Score

### 8.5. Suppl 5. MISS□HF

### 8.6. Suppl 6. ROC for MISS-HP-Persian

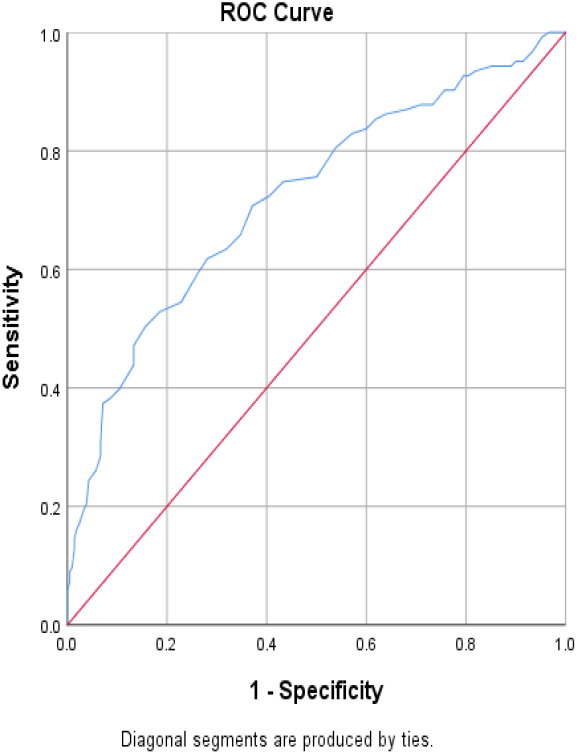

